# Disability disparities in STEM: Gaps in salaries and representation for doctorate recipients with disabilities in the U.S., 2019

**DOI:** 10.1101/2022.12.04.22283081

**Authors:** Franz Castro, Elizabeth Stuart, Jennifer Deal, Varshini Varadaraj, Bonnielin K. Swenor

## Abstract

**Introduction:** There is paucity of data examining disparities in salary and representation for disabled scientists, which is needed to advance inclusion and equity for people with disabilities in Science, Technology, Engineering and Mathematics (STEM).

**Methods:** Analyses used cross-sectional data from the 2019 Survey of Doctorate Recipients. We compared salaries between doctorate recipients with and without disabilities who were currently employed in STEM (N = 704,013), but who were otherwise similar on socioeconomic, degree and job-related characteristics, using propensity score weighting to carry out balanced comparisons, and further examined these differences in salary in the subset of doctorate recipients working in STEM at academic institutions (N = 219,413). In the subset of participants working in academia, we examined whether the representation of people with disabilities differed across categories of academic career milestones using chi-square tests (α = 0.05).

**Results:** Doctorate recipients working in STEM with early onset disabilities (identified <25 years of age) earned $10,580 less per year than non-disabled workers, and in the subset of academic workers this difference was larger (-$14,360). Salaries appeared lower for people with late onset disabilities as compared to those without, although these differences did not reach statistical significance. We observed an underrepresentation of academics with disabilities at higher faculty ranks (p<0.0001), among Deans/Presidents (p<0.0001) and among those with tenure (p: 0.0004).

**Conclusion:** These findings support a need to expand efforts to foster inclusion, provide equal opportunities for career advancement, and improve working conditions for people with disabilities in STEM.

## Introduction

Although over a quarter of U.S. adults have a disability, people with disabilities account for only 10% of the science, technology, engineering, and mathematic (STEM) workforce.(1, 2) Disability inclusion in STEM is critical to the diversification of sciences,(3) and a matter of both economic development and equity. Engaging the full range of diverse talent in STEM leads to gains for the country’s scientific landscape, as research and innovation are strengthened when people with diverse life experiences and knowledge contribute to the solution of complex problems.(4)

Salary is an important metric of equity in the workforce. Historical wage gaps by gender and race have shed light on systematic biases impacting workers from marginalized groups.(5, 6) Consequently, equal pay has been at the center of legislation and policy discussion for decades. Previous reports have highlighted that in science and engineering fields, women and underrepresented racial minorities earn on average $25,000 and $15,000 less per year as compared to their male and White colleagues, respectively.(2) However, there is paucity of data examining disparities in salary and representation for disabled scientists, which is needed to advance inclusion and equity for people with disabilities in STEM. To fill these data gaps, we used data from the 2019 Survey of Doctorate Recipients (SDR),(7) containing data on 80,882 participants, representing 1,148,817 U.S. research doctorate degree recipients with their STEM degrees awarded between 1973 and 2017, regardless of whether they were still active in the workforce or not.

This study aimed to compare salaries between doctorate recipients with and without disabilities who were currently employed in STEM, but who were otherwise similar on socioeconomic, degree and job-related characteristics, using propensity score weighting to carry out balanced comparisons. We further examined these differences in salary in the subset of doctorate recipients working in STEM at academic institutions. Our secondary objective was limited to the subset of participants working in academia, and examined whether the representation of people with disabilities differed across categories of academic career milestones.

## Materials and Methods

Analyses used cross-sectional data from the 2019 Survey of Doctorate Recipients (SDR),(7) a biennial survey conducted by the National Science Foundation (NSF) since 1973. The SDR 2019 collects demographic, educational, and career history information from individuals that obtained a U.S. research doctorate degree between 1973 and 2017 in a science, engineering, or health field. Research doctorate degrees are defined by the SDR as requiring completion of an original intellectual contribution or a dissertation and are not primarily intended for the practice of a profession. Therefore, recipients of professional doctorate degrees were not included in the survey (e.g., MD, DPharm, PsyD, etc.). The NSF’s definition of science, technology, engineering, and mathematics (STEM) fields extends beyond natural sciences, mathematics, engineering, computer and information sciences, and includes behavioral sciences such as psychology, economics, sociology, and political science.(8)

The SDR relies on a fixed panel design, adding to the study a sample of 10,000 new graduates every other year. Subjects who graduated between 1973 and 2017 were sampled from the Doctorate Records File (DRF), a yearly census of all doctorate recipients in the U.S, regardless of whether they had dropped out of the labor market or not. For the 2019 SDR, new graduates had their degrees awarded between July 2015 and June 2017, for a total sample size of 120,000 participants, and 80,882 respondents (response rate: 69%). Excluded from the SDR were participants that were 76 years of age or older, institutionalized, or terminally ill as of February 1^st^, 2019, resulting in a weighted total of 1,148,817 doctorate recipients. The SDR utilizes survey weights that account for differential sampling rates, adjustments for nonresponse, unknown eligibility, and for aligning the sample with the DRF distribution on gender, race/ethnicity, location, degree field, and year. Prior publications have described the survey methodology in greater detail.(7) There were no missing data for the variables used in the main analyses.

Our primary analyses were restricted to SDR participants from 2019 currently employed in a STEM field, living in the U.S., and working for an employer based in the U.S. at the time of the survey (N = 704,013). In secondary analyses, we further restricted the study population to participants working at academic institutions (N = 219,413, 31.2% of the total study population), defined as postsecondary educational institutions (4-year colleges or universities, medical schools, university-affiliated research institutes) where the following were available: faculty ranks, tenure, and academic positions (e.g. Deans or Presidents). There were no observations with missing data for the variables used in the analyses.

### Definitions of salary and disability

Participants were asked about their basic annual salaries on their principal job, before tax deductions, and about the usual degree of difficulty (none, slight, moderate, severe, or unable to do) they experienced in any of the following domains: 1) seeing words or letters in ordinary newsprint, 2) hearing what is normally said in conversation with another person, 3) walking without human or mechanical assistance or using stairs, 4) lifting or carrying something as heavy as 10 pounds, or 5) concentrating, remembering, or making decisions because of a physical, mental, or emotional condition. Those who responded “moderate”, “severe”, or “unable to do” for any activity were classified as having a disability.

To better characterize the impact of age of disability onset on career outcomes, we used the following disability categories: no disability, disability first identified at 25 years of age or later (hereafter referred to as late onset disability), and disability first identified before 25 years of age (hereafter referred to as early onset disability). Since 44.1% of all doctorate recipients earn their degrees between 26 and 30 years of age, and 0.6% before 26 years of age,(9) we chose 25 years as a threshold for early and late disability to allow all subjects to have a non-zero probability of being in a given exposure group.

### Socioeconomic, degree, job-related, and academic career covariates

We examined the distribution of covariates of interest across groups, such as sex (male, female), age (<35 35-44, 45-54. 55-64, ≥65 years), race/ethnicity (Non-Hispanic White, Hispanic, Black, Asian, or Other [including multiracial]), marital status (never married, separated or divorced, widowed, married or living in a marriage-like relationship), field of doctoral degree (biological, agricultural, and environmental life sciences; computer and information sciences; mathematics and statistics; physical sciences, geosciences, atmospheric, and ocean sciences; psychology; social sciences; engineering; health), job field (computer and mathematical sciences; biological, agricultural, and other life sciences; physical and related sciences; social and related sciences; engineering; other occupations related to science and engineering), region of the U.S. where the employer is located (West, Midwest, Northeast, South), having a full-time position throughout the entire year (defined as working at least 36 hours per week and 52 weeks per year), and working at an academic institution. The following covariates were further examined only for participants working at academic institutions: faculty rank (Instructor or Lecturer, Assistant Professor, Associate Professor, Professor), tenure status (not on tenure-track and not tenured, on tenure-track, tenured), and being a Dean or a President at their institution.

### Rationale for using propensity scores

Propensity scores are commonly used in non-experimental studies of treatments, exposures, or interventions to adjust for confounders by using the propensity score to match, weight, or subclassify the groups and thus create covariate “balance” – similarity – across the exposure groups.(10) In the propensity score adjusted samples (e.g., the matched or weighted samples), the distribution of observed covariates should be similar between groups. In our application we are using propensity score not for causal effect estimation, but rather for this covariate balancing property, to create “balanced comparisons” in which we can compare salaries between doctorate recipients with and without disabilities who were otherwise similar on socioeconomic, degree field, and job-related characteristics.

### Propensity score computation using generalized boosted models

Propensity scores were computed using generalized boosted models (GBM), a machine learning approach that predicts a dichotomous binary treatment indicator using many simple regression trees iteratively combined to fit an overall piecewise constant function in a flexible way. In this scenario of multiple exposure groups (no disability, late, and early onset disability), dummy indicators for each of the disability groups are created and separate GBMs are fitted to each dummy treatment indicator.(11) When computing the propensity scores, we included factors that were relevant predictors of salary: race, sex, marital status, field of doctoral degree, job field, region of the U.S. where the employer is located, receiving federal funding, and having a full-time job. Propensity scores were computed across strata of age (≤34 years, 35-44 years, 45-54 years, 55-64 years, ≥65 years) to partially remove the effect of longevity in the workforce.

### Inverse probability of treatment weighting (IPTW) and examination of covariate balance

After the propensity scores were estimated using GBM they were used to create weights to weight individuals with late, early onset, and without disabilities to be similar to one another. In particular, Inverse Probability of Treatment Weighting (IPTW) was used. IPTW gives each subject a weight based on the propensity score to create a synthetic sample in which the distribution of observed covariates is independent of disability status (at least with respect to the observed characteristics) and therefore similar between groups. A subject’s weight is equal to the inverse of the probability of being in their group (e.g., for individuals without disabilities, one over their probability of not having a disability, as estimated from the propensity score model given the observed covariates), and constitutes a form of model-based direct standardization.(10) We assessed covariate balance using standardized mean differences, and values below 0.1 were used to determine that a good covariate balance was achieved. IPTW weights were multiplied by survey weights to account for the complex survey design.

### Linear regression, association between disability and salary

We conducted balanced comparisons on salaries between a) doctorate recipients without disabilities and those with late onset disabilities, and b) between those without disabilities and those with early onset disabilities. Using a doubly robust approach, the regression models adjusted for the same set of covariates as used in the propensity score models.(12) By using linear regression with sandwich variance estimators, we calculated beta coefficients (β) with 95% confidence intervals (CIs) for the associations between disability and salary. Besides combining IPTW weights with survey weights when balancing groups, regression models additionally accounted for survey weights.(13) A similar analysis was conducted for doctorate recipients working in STEM at academic institutions, for which additional covariates were added to the propensity score computation and outcome regression model: faculty rank, tenure status, and whether the participant was a Dean or a President at their institution.

### Representation of doctorate recipients with disabilities across categories of academic career milestones

We aimed at better understanding how people with disabilities are represented in different categories of achievement of academic career milestones. In the subset of STEM workers at academic institutions (N=219,413), we examined the proportions (survey-weighted and age-standardized using the 2010 U.S. Census population) of participants without, with late and early onset disabilities across the following categories:(14) 1) faculty rank (Instructor/Lecturer, Assistant Professor, Associate Professor, Professor), 2) tenure status (not tenured, on tenure track, tenured), 3) being a Dean/President (no, yes), 4) receiving federal funding for their work/research (no, yes). For each category, we tested differences using chi-square tests (α = 0.05). We conducted supplementary analyses showing the representation of people with disabilities among workers who received federal funding (grants or contracts), by type of U.S. agency.

SDR 2019 data is publicly available on the NSF’s website (https://ncsesdata.nsf.gov/datadownload/), and Institutional Review Board approval was not required since data were de-identified. All analyses were conducted using R Studio, version 2021.09.1. Documentation on the *weightit* and *cobalt* R packages used for propensity score computation and assessment of covariate balance can be found at https://cran.r-project.org/web/packages/cobalt/index.html and https://cran.r-project.org/web/packages/WeightIt/index.html

## Results

Out of the 704,013 doctorate recipients working in STEM, 646,652 (91.9%, 95% CI: 91.5%, 92.2%) did not report disabilities, 36,807 (5.2%, 95% CI: 5.0%, 5.5%) reported late onset disabilities, and 20,554 (2.9%, 95% CI: 2.7%, 3.1%) reported early onset disabilities **(Fig 1, S1 Fig)**. There was a lower proportion of female doctorate recipients as compared to males across all three study groups. The highest representation of female STEM workers was observed in the no disability group (35.6% [95% CI: 35.0%, 36.2%]), and the lowest in the late onset disability group (29.5% [95% CI: 27.4%, 31.8%]). The age distributions were fairly similar between the no disability and the early onset disability groups (mostly 35-39 years), whereas in the late onset disability group more than half of the participants were 55 years of age and older. Although participants in the early onset disability group were younger overall, the percentage of participants aged 45-49 and 50-54 years was similar across the three study groups (**Table 1**).

**Table 1.** Characteristics of doctorate recipients working in STEM, and covariate balance for propensity score weighting analysis.

| Variable | No disability, %<br>(95% CI)<br>(N = 646,652) | Late onset<br>disability (first<br>identified at 25<br>years of age or<br>later), % (95%<br>CI)<br>(N = 36,807) | Early onset<br>disability (first<br>identified before<br>25 years of age),<br>% (95% CI)<br>(N = 20,554) | Covariate balance after<br>propensity score weighting<br>(sample: all STEM workers) |  | Covariate balance after<br>propensity score weighting<br>(sample: STEM workers at<br>academic institutions) |  |
| --- | --- | --- | --- | --- | --- | --- | --- |
|  |  |  |  | SMD<br>(No<br>disability vs.<br>Late onset<br>disabilities)<br>a,b | SMD<br>(No<br>disability vs.<br>Early onset<br>disabilities)<br>a,b | SMD<br>(No<br>disability vs.<br>Late onset<br>disabilities)<br>a,b | SMD<br>(No<br>disability vs.<br>Early onset<br>disabilities)<br>a,b |
| Sex |  |  |  |  |  |  |  |
| Female <sup>c</sup> | 35.6 (35.0, 36.2) | 29.5 (27.4, 31.8) | 34.4 (31.3, 37.5) | -0.0078 | 0.0017 | -0.008 | 0.027 |
| Male | 64.4 (63.8, 65) | 70.5 (68.2, 72.6) | 65.6 (62.5, 68.7) | 0.0078 | -0.0017 | 0.008 | -0.027 |
| Age (years) |  |  |  |  |  |  |  |
| ≤29 <sup>c</sup> | 1.1 (1.0, 1.3) | 0.1 (0.0, 0.2) | 1.8 (1.2, 2.8) | -0.0065 | 0.0014 | -0.005 | -0.003 |
| 30-34 | 12.1 (11.7, 12.5) | 2.5 (1.8, 3.5) | 13.3 (11.3, 15.4) | -0.0093 | 0.0114 | -0.062 | -0.04 |
| 35-39 | 16.6 (16.2, 17.1) | 5.8 (4.7, 7.1) | 19.2 (16.5, 22.1) | -0.0055 | 0.0083 | -0.048 | 0.014 |
| 40-44 | 14.7 (14.2, 15.1) | 7.8 (6.4, 9.5) | 14.5 (12.2, 17.2) | -0.0093 | 0.0061 | -0.014 | 0.001 |
| 45-49 | 12.4 (12.0, 12.8) | 12.6 (11.0, 14.5) | 13.5 (11.2, 16.1) | 0.0037 | -0.0106 | 0.028 | -0.003 |
| 50-54 | 11.1 (10.7, 11.5) | 13.4 (11.7, 15.2) | 10.0 (8.3, 12.1) | 0.0003 | -0.0043 | 0.029 | 0.042 |
| 55-59 | 11.1 (10.7, 11.5) | 16.4 (14.6, 18.5) | 10.3 (8.3, 12.7) | 0.0073 | -0.0039 | 0.025 | 0.003 |
| 60-64 | 9.7 (9.3, 10.1) | 15.4 (13.5, 17.4) | 7.7 (6.0, 9.7) | 0.0068 | -0.0036 | 0.021 | 0.001 |
| 65-69 | 7.2 (6.8, 7.5) | 14.6 (12.9, 16.6) | 5.3 (4.1, 6.8) | 0.0072 | -0.0048 | 0.017 | -0.017 |
| 70-75 | 4.1 (3.8, 4.3) | 11.4 (9.9, 13.2) | 4.5 (3.1, 6.3) | 0.0052 | -0.0002 | 0.01 | 0.002 |
| Race/ethnicity |  |  |  |  |  |  |  |
| White only, NH <sup>c</sup> | 64.2 (63.6, 64.8) | 69.6 (67.1, 72.0) | 67.9 (64.5, 71.1) | 0.0185 | 0.0154 | 0.028 | 0.071 |
| Hispanic | 4.4 (4.2, 4.6) | 4.3 (3.6, 5.2) | 5.2 (4.2, 6.3) | 0.0024 | 0.0069 | -0.004 | 0.006 |
| Black only, NH | 3.3 (3.2, 3.5) | 2.9 (2.3, 3.6) | 3.3 (2.3, 4.5) | -0.0062 | -0.001 | -0.009 | 0.003 |
| Asian only, NH | 26.6 (26.0, 27.2) | 21.1 (19.0, 23.5) | 21.6 (18.6, 24.9) | -0.012 | -0.0205 | -0.016 | -0.08 |
| Other, including<br>multiracial, NH | 1.4 (1.3, 1.6) | 2.1 (1.4, 3.0) | 2.1 (1.3, 3.3) | -0.0027 | -0.0008 | 0.001 | -0.001 |
| Marital status |  |  |  |  |  |  |  |
| Never married <sup>c</sup> | 9.7 (9.4, 10.1) | 7.7 (6.4, 9.1) | 14.5 (12.2, 17.2) | -0.0046 | -0.0082 | -0.01 | -0.013 |
| Separated,<br>Divorced | 5.8 (5.6, 6.2) | 9.9 (8.5, 11.5) | 7.2 (5.7, 9.0) | -0.0027 | -0.0076 | -0.006 | -0.012 |
| Widowed | 0.8 (0.7, 0.9) | 1.7 (1.2, 2.5) | 0.7 (0.3, 1.6) | -0.0015 | -0.0069 | -0.001 | -0.008 |
| Married or living<br>in a marriage-like<br>relationship | 83.6 (83.1, 84.1) | 80.7 (78.6, 82.6) | 77.6 (74.5, 80.3) | 0.0087 | 0.0227 | 0.018 | 0.033 |
| Degree field |  |  |  |  |  |  |  |
| Biological, agricultural, and environmental life sciences <sup>c</sup> | 26.1 (25.6, 26.7) | 23.4 (21.4, 25.7) | 26.8 (24, 29.9) | 0.0129 | 0.0212 | 0.017 | 0.011 |
| Computer and information sciences | 3.9 (3.7, 4.2) | 2.8 (2.1, 3.7) | 3.9 (2.7, 5.7) | -0.0065 | -0.0072 | -0.004 | 0.0 |
| Mathematics and statistics | 4.6 (4.4, 4.8) | 4.6 (3.7, 5.7) | 4.5 (3.4, 6.1) | -0.0012 | -0.0049 | 0.0 | -0.012 |
| Physical sciences, geosciences, atmospheric, and ocean sciences | 16.2 (15.8, 16.7) | 15.8 (14.0, 17.9) | 16.2 (13.9, 18.8) | 0.0033 | -0.0089 | -0.011 | 0.001 |
| Psychology | 12.8 (12.4, 13.2) | 14.4 (12.7, 16.4) | 13.5 (11.3, 16.1) | -0.0022 | -0.0027 | -0.013 | -0.007 |
| Social sciences | 9.8 (9.5, 10.2) | 13.8 (12.2, 15.7) | 11.8 (9.8, 14.1) | -0.0004 | 0.0044 | 0.012 | 0.025 |
| Engineering | 22.0 (21.4, 22.5) | 19.3 (17.1, 21.7) | 18.4 (15.6, 21.5) | -0.0028 | -0.0036 | 0.002 | -0.022 |
| Health | 4.6 (4.3, 4.8) | 5.8 (4.7, 7.1) | 4.9 (3.8, 6.3) | -0.0031 | 0.0016 | -0.005 | 0.004 |
| Job field |  |  |  |  |  |  |  |
| Computer and mathematical sciences <sup>c</sup> | 15.0 (14.5, 15.4) | 12.0 (10.4, 13.6) | 14.7 (12.3, 17.4) | -0.0059 | -0.0134 | -0.001 | -0.015 |
| Biological, agricultural, and other life sciences | 22.2 (21.7, 22.7) | 20.6 (18.6, 22.7) | 25.0 (22.3, 28.0) | 0.0125 | 0.015 | 0.022 | 0.018 |
| Physical and related sciences | 11.9 (11.6, 12.3) | 12.2 (10.6, 14.0) | 12.6 (10.6, 14.9) | -0.0054 | 0.0032 | -0.011 | 0.004 |
| Social and related sciences | 20.5 (20.0, 21.1) | 25.4 (23.1, 27.7) | 22.5 (19.7, 25.5) | 0.003 | 0.0023 | -0.001 | 0.009 |
| Engineering | 17.2 (16.7, 17.7) | 16.3 (14.3, 18.6) | 14.7 (12.3, 17.5) | 0.0009 | 0.0024 | 0.003 | -0.012 |
| Other occupations related to science and engineering | 13.1 (12.7, 13.5) | 13.6 (11.9, 15.5) | 10.5 (8.7, 12.6) | -0.0051 | -0.0095 | -0.012 | -0.003 |
| Region of the U.S. where employer is located |  |  |  |  |  |  |  |
| West <sup>c</sup> | 29.5 (29.0, 30.1) | 26.1 (23.9, 28.5) | 28.4 (25.4, 31.6) | -0.0036 | 0.0093 | 0.015 | -0.006 |
| Midwest | 17.5 (17.0, 17.9) | 16.9 (15.0, 18.9) | 19.8 (17.1, 22.7) | 0.0075 | -0.0051 | -0.007 | -0.007 |
| Northeast | 22.6 (22.0, 23.1) | 19.9 (17.9, 22.0) | 23.0 (20.2, 26.0) | -0.0124 | -0.0159 | -0.006 | 0.01 |
| South | 30.4 (29.9, 31.0) | 37.2 (34.6, 39.8) | 28.9 (25.9, 32.0) | 0.0085 | 0.0117 | -0.002 | 0.003 |
| Works full time |  |  |  |  |  |  |  |
| No <sup>c</sup> | 30.8 (30.3, 31.4) | 42.9 (40.3, 45.6) | 33.9 (30.6, 37.3) | 0.0099 | -0.0099 | 0.044 | 0.027 |
| Yes | 69.2 (68.6, 69.7) | 57.1 (54.4, 59.7) | 66.1 (62.7, 69.4) | -0.0099 | 0.0099 | -0.044 | -0.027 |
| Receives federal funding |  |  |  |  |  |  |  |
| No <sup>c</sup> | 72.1 (71.5, 72.6) | 72.9 (70.5, 75.2) | 73.7 (70.7, 76.5) | -0.0072 | 0.01 | -0.022 | 0.009 |
| Yes | 27.9 (27.4, 28.5) | 27.1 (24.8, 29.5) | 26.3 (23.5, 29.3) | 0.0072 | -0.01 | 0.022 | -0.009 |
| Works at an academic institution <sup>e</sup> |  |  |  |  |  |  |  |
| No <sup>c</sup> | 57.7 (57.1, 58.3) | 51.6 (48.9, 54.2) | 52.5 (49.1, 55.9) | - | - | - | - |
| Yes | 42.3 (41.7, 42.9) | 48.4 (45.8, 51.1) | 47.5 (44.1, 50.9) | - | - | - | - |
| Faculty rank <sup>f</sup> |  |  |  |  |  |  |  |
| Instructor/Lecturer <sup>c</sup> | 7.7 (7.2, 8.3) | 7.7 (5.8, 10.2) | 7.0 (4.9, 9.9) | - | - | -0.014 | -0.023 |
| Assistant Professor | 26.4 (25.5, 27.3) | 14.8 (12.2, 17.8) | 33.7 (28.7, 39.1) | - | - | -0.081 | -0.017 |
| Associate Professor | 27.7 (26.8, 28.6) | 24.8 (21.5, 28.5) | 32.0 (26.7, 37.7) | - | - | 0.016 | 0.03 |
| Professor | 38.2 (37.2, 39.2) | 52.7 (48.7, 56.6) | 27.3 (22.7, 32.5) | - | - | 0.08 | 0.01 |
| Tenure status <sup>f</sup> |  |  |  |  |  |  |  |
| Not on tenure track <sup>c</sup> | 20.0 (19.1, 20.8) | 15.2 (12.5, 18.4) | 19.8 (16.0, 24.3) | - | - | -0.031 | -0.038 |
| On tenure-track but not tenured | 20.8 (20.0, 21.7) | 11.5 (9.2, 14.1) | 27.0 (22.3, 32.4) | - | - | -0.065 | -0.006 |
| Tenured | 59.2 (58.2, 60.2) | 73.3 (69.6, 76.7) | 53.2 (47.4, 58.8) | - | - | 0.096 | 0.044 |
| Dean or President<sup>f</sup> |  |  |  |  |  |  |  |
| No <sup>c</sup> | 92.0 (91.5, 92.5) | 91.9 (89.8, 93.6) | 94.6 (92.1, 96.4) | - | - | 0.003 | 0.005 |
| Yes | 8.0 (7.5, 8.5) | 8.1 (6.4, 10.2) | 5.4 (3.6, 7.9) | - | - | -0.003 | -0.005 |

1 a The standardized differences in means between the no disability and each disability group were calculated as difference in means between groups divided by the SD of the 2 disability group for each of the factors included in the propensity score computation, expressed numerically.
3 b Propensity scores were computed stratifying across strata of age ( $\leq 34$ years, 35-44 years, 45-54 years, 55-64 years, $\geq 65$ years).
4 c This group was used as the reference group for the other levels of the same factor.
6 e Not used in computation of propensity scores
7 f Only for those working in academic institutions
NH: Non-Hispanic.
SMD: Standardized mean difference

**Fig 1.**
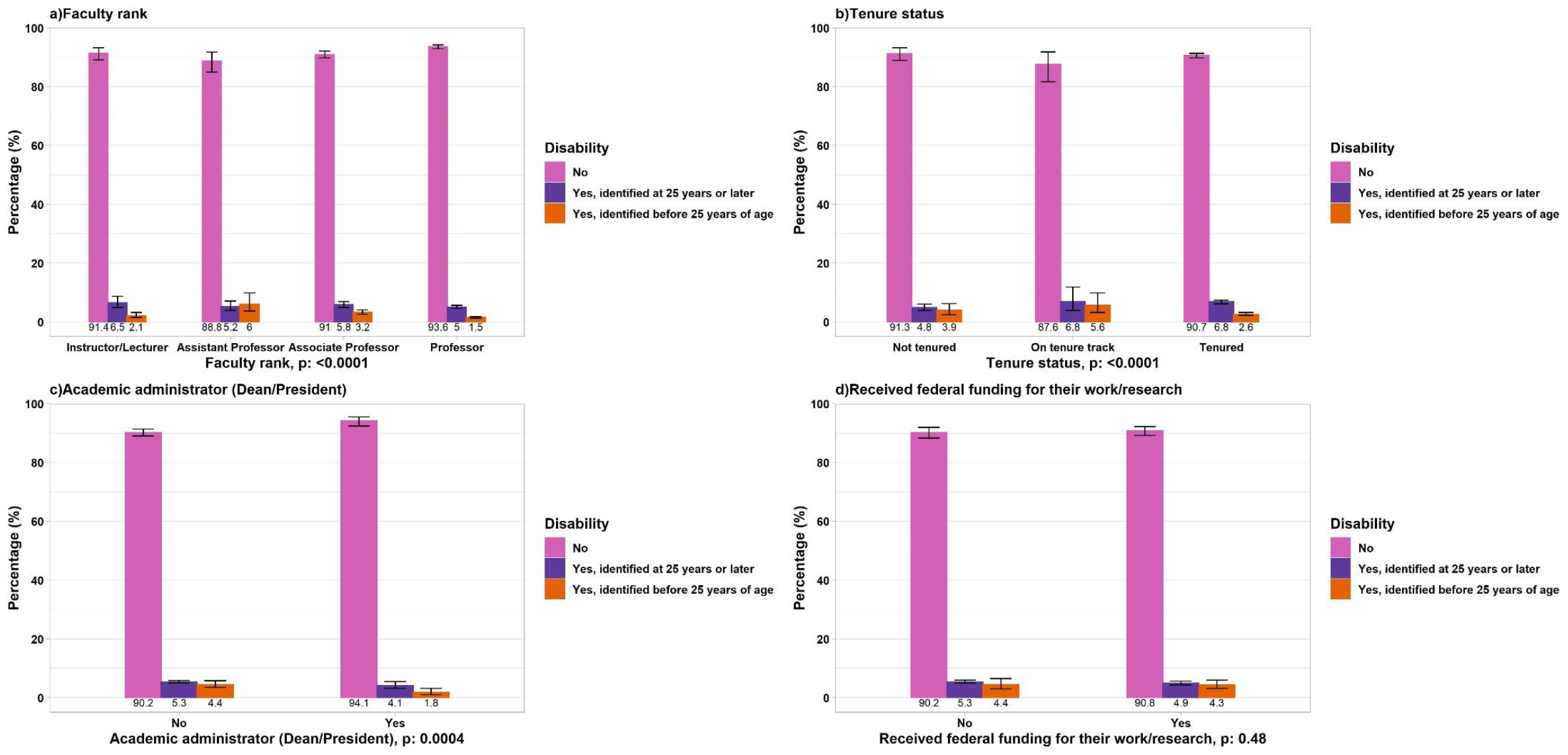
Representation (survey-weighted, age-standardized proportions) of doctorate recipients with disabilities working in STEM across categories of academic career milestones. a) Faculty rank, b) Tenure status, c) Academic administrator (Dean/President), d)Received federal funding for their work/research. Other: Participants identified as “Other races including multiracial individuals, non-Hispanic”. For each panel, the total denominator is the number of doctorate recipients working in STEM at academic institutions (N=219,413).

The late onset disability group had the lowest representation of Hispanic and Black doctorate recipients (4.3% [95% CI: 3.6%, 5.2%] and 2.9% [95% CI: 2.3%, 3.6%]), as compared to the early onset disability group with 5.2% (95% CI: 4.2%, 6.3%) and 3.3% (95% CI: 2.3%, 4.5%), respectively. Overall, the largest percentage of doctorate degrees were awarded in the field of biological/agricultural/environmental life sciences, and the lowest percentage in the field of computer/information sciences. For participants without disabilities, 22.0% (95% CI: 21.4%, 22.5%) of the degrees were awarded in the field of engineering, in comparison to 19.3% (95% CI: 17.1%, 21.7%) in the late onset disability group, and 18.4% (95% CI: 15.6%, 21.5%) among those with early disabilities. Conversely, participants with disabilities had a higher percentage of degrees in the social sciences (13.8% [95% CI: 12.2%, 15.7%] and 11.8% [95% CI: 9.8%, 14.1%], respectively), as compared to those without disabilities (9.8% [95% CI: 9.5%, 10.2%]). The proportion of full-time workers (versus part-time) was higher in the no disability group (69.2% [95% CI: 68.6%, 69.7%]) than in the early (66.1% [95% CI: 62.7%, 69.4%]) and late (57.1% [95% CI: 54.4%, 59.7%]) onset disabilities groups (**Table 1**). Among STEM workers without disabilities, 27.9% (95% CI: 27.4%, 28.5%) reported receiving federal funding (e.g. contracts, grants) to support their work, versus 26.3% (95% CI: 23.5%, 29.3%) in the early onset disability group.

A total of 219,413 doctorate recipients were working at academic institutions, of whom 198,689 (90.6%, 95% CI: 89.9%, 91.0%) had no disabilities, and 13,929 (6.4%, 95% CI: 5.9%, 7.0%) and 6,794 (3.1%, 95% CI: 2.8%, 3.5%) had late and early onset disabilities, respectively. While most participants without disabilities and with late onset disabilities reported being Professors (38.2% [95% CI: 37.2%, 39.2%] and 52.7% [95% CI: 48.7%, 56.6%]), the largest percentage of those with early onset disabilities reported being Assistant Professors (33.7% [95% CI: 28.7%, 39.1%]). In fact, among those with early onset disabilities, only 27.3% (95% CI: 22.7%, 32.5%) were Professors. The proportion of doctorate recipients reporting being Deans or Presidents was highest among those with late onset disabilities (8.1% [95% CI: 6.4%, 10.2%]) and those without disabilities (8.0% [95% CI: 7.5%, 8.5%]), and lowest for those with early onset disabilities (5.4% [95% CI: 3.6%, 7.9%]). We observed similar results for tenure attainment: 73.3% (95% CI: 69.6%, 76.7%) of doctorate recipients with late onset disabilities and 59.2% (95% CI: 58.2%, 60.2%) of those without disabilities were tenured, as compared to 53.2% (95% CI: 47.4%, 58.8%) in the early onset disability group.

We used propensity score weighting to address covariate imbalance in the data. Propensity scores represent the probability of being in an exposure group (no disability, late onset, and early onset disability) given a set covariates.(10) Therefore, conditional on the propensity scores, the distribution of observed covariates will be similar between groups. This approach applies a calculated propensity score weight to each participant, equal to the inverse of their propensity score, as a form of direct standardization. Additionally, we stratified by age (≤34 years, 35-44 years, 45-54 years, 55-64 years, ≥65 years) to account for the effect of longevity in the workforce, and propensity score weights were multiplied by survey weights to account for the survey design.

**Table 2** summarizes the results of the linear regression model for comparing salaries after using propensity score weighting to balance groups on socioeconomic (sex, age, race/ethnicity, marital status), degree field, and job-related characteristics (job field, region of the U.S. where the employer is located, receiving federal funding, having a full-time job). For doctorate recipients working in STEM, having a disability first identified before 25 years of age was associated with a significantly lower annual salary in comparison to their non-disabled peers (β: -$10,580, 95% CI: -$13,661, -$7,499; p: <0.0001). These differences were larger in the subset of STEM workers in academic institutions, with those with an early onset disability having a salary $14,360 lower than those without disabilities (β: -$14,360; 95% CI: -$17,546, -$11,175; p: <0.0001) (**Table 3**). Salaries appeared lower for people with late onset disabilities as compared to those without, although these differences did not reach statistical significance for the overall STEM workforce (β: -$7,577; 95% CI: -$15,452, $299; p: 0.06), or for the subset of academic STEM workers (β: - $3,976; 95% CI:-$11,607, $3,656; p: 0.31) (**Tables 2 and 3**). **S2 Fig** and **S3 Fig** show a summary of the covariate balance for both samples (all STEM workers and STEM workers at academic institutions).

**Table 2.**
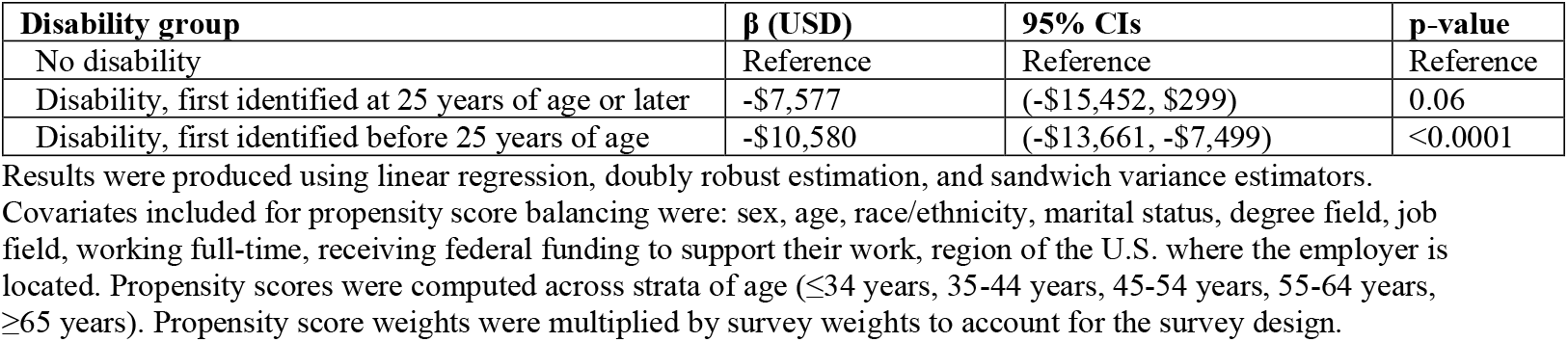
Association between disability and salary for doctorate recipients working in STEM using propensity score weighting, linear regression.

| Disability group | $\beta$ (USD) | 95% CIs | p-value |
| --- | --- | --- | --- |
| No disability | Reference | Reference | Reference |
| Disability, first identified at 25 years of age or later | -\$7,577 | (-\$15,452, \$299) | 0.06 |
| Disability, first identified before 25 years of age | -\$10,580 | (-\$13,661, -\$7,499) | <0.0001 |

Results were produced using linear regression, doubly robust estimation, and sandwich variance estimators. Covariates included for propensity score balancing were: sex, age, race/ethnicity, marital status, degree field, job field, working full-time, receiving federal funding to support their work, region of the U.S. where the employer is located. Propensity scores were computed across strata of age ( $\leq 34$ years, 35-44 years, 45-54 years, 55-64 years, $\geq 65$ years). Propensity score weights were multiplied by survey weights to account for the survey design.

**Table 3.** Association between disability and salary for doctorate recipients working in STEM at academic institutions using propensity score weighting, linear regression.

| Disability group | Coefficient (USD) | 95% CIs | p-value |
| --- | --- | --- | --- |
| No disability | Reference | Reference | Reference |
| Disability, first identified at 25 years of age or later | -\$3,976 | (-\$11,607, \$3,656) | 0.31 |
| Disability, first identified before 25 years of age | -\$14,360 | (-\$17,546, -\$11,175) | <0.0001 |

Results were produced using linear regression, doubly robust estimation, and sandwich variance estimators. Covariates included for propensity score balancing were: sex, age, race/ethnicity, marital status, degree field, job field, working full-time, receiving federal funding to support their work, region of the U.S. where the employer is located, tenure status, faculty rank, being an academic administrator (Dean/President). Propensity scores were computed across strata of age ( $\leq 34$ years, 35-44 years, 45-54 years, 55-64 years, $\geq 65$ years). Propensity score weights were multiplied by survey weights to account for the survey design.

In the subset of doctorate recipients working in academia, we investigated the representation of people with disabilities in categories of academic career milestones (faculty rank, tenure status, being a Dean/President, receiving federal funding for their work/research). When examining the representation of people with disabilities among different categories of faculty rank, we observed that among doctorate recipients with the rank of Professor, only 5.0% (95% CI: 4.5%, 5.6%) had late onset and 1.5% (95% CI: 1.2%, 1.8%) early onset disabilities. Conversely, among Instructors/Lecturers, the representation of people with late and early onset disabilities was 6.5% (95% CI: 4.8%, 8.7%) and 2.1% (95% CI: 1.5%, 3.1%) (p < 0.0001, chi-square test). Among tenured STEM professionals, the percentages of people with disabilities identified later and earlier in life were 6.8% (95% CI: 6.1%, 7.4%) and 2.6%, (95% CI: 2.2%, 3.1%), as compared to 4.8% (95% CI: 3.8%, 6.0%) and 3.9% (95% CI: 2.4%, 6.2%) among those who were not tenured (p < 0.0001). The percentage of participants with late and early onset disabilities among Deans/Presidents was lower (4.1% [95% CI: 3.1%, 5.4%] and 1.8% [95% CI: 1.0%, 3.1%], respectively) than among those who were not Deans/Presidents (5.3% [95% CI: 4.9%, 5.8%] and 4.4% [95% CI: 3.4%, 5.7%]) (p: 0.0004). We observed similar proportions of people with disabilities among those who did and did not receive federal funding to support their work/research (p: 0.48) (**Fig 1**).Among STEM workers receiving federal funding (**S4 Fig**), the highest representation of people with late onset disabilities was observed among STEM workers funded by the Department of Education (6.6% [95% CI: 3.8%, 11.1%]), and the lowest among those funded by the National Aeronautics and Space Administration (NASA, 4.3% [95% CI: 2.8%, 6.5%]). Conversely, the highest representation of STEM workers with early onset disabilities was among those supported by the Department of Defense (5.8% [95% CI: 2.6%, 12.3%]), and the lowest for those funded by NASA (1.9% [95% CI: 1.1%, 3.2%]).

## Discussion

This study of 704,013 doctorate recipients in the U.S. highlights lower salaries for STEM workers with disabilities and underrepresentation in academic leadership positions, both in the overall STEM workforce and in the subset of those working at academic institutions. When using propensity score weighting to compare groups by disability status but who were similar on socioeconomic and job-related characteristics, we observed that doctorate recipients with early onset disabilities had annual salaries that were on average $10,580 lower than their non-disabled counterparts, and that this difference was larger in the subset of STEM workers in academia (-$14,360, early onset versus no disability). Salaries appeared lower in the late onset disability than in the no disability group, although these differences were not statistically significant. We found an underrepresentation of doctorate recipients with disabilities among Professors, tenured academics, and Deans/Presidents.

Prior to the propensity score weighting, the group of doctorate recipients with late onset disabilities was older and had higher proportions of Professors and tenured workers when compared to both the no disability and the early onset disability groups. Since more than half of STEM workers with late onset disabilities were 55 years of age and older, it is possible that this group was mostly comprised of workers with disabilities related to aging. Furthermore, three quarters of participants in this group were tenured, and 54% reported being Professors, a proportion twice as high as that observed for people with disabilities identified before 25 years of age. These differences between groups based on age of disability onset might have been driven by the fact that adults with disabilities acquired early in life face a unique set of challenges for entering the STEM workforce, for earning equal wages, and for accessing equal career advancement opportunities as compared to those with late onset disabilities or without disabilities.(15) Regardless of when their disabilities were identified, the disability community faces a two-sided attrition in their career trajectories in STEM: while younger individuals with disabilities are impacted by structural barriers to ultimately securing STEM education and jobs, older workers with disabilities face the pressures of retirement due to working conditions characterized by lack of accessibility in the physical environment and institutional policies that hinder the procurement of accommodations.(16)

Previous publications have shown that a household containing an adult with a disability requires 28% more income to obtain the same standard of living as a similar household without a member with a disability.(17) Despite this, a report by the National Science Foundation shows that scientists and engineers with at least one disability have an unemployment rate higher than that for the overall U.S. labor force (5.3% versus 3.7%), and that doctorate recipients with any disability working in STEM earn $9,000 less per year than their non-disabled counterparts.(2) Salary gaps for workers with disabilities emerge early in their careers,(15, 18) when entry-level wages and work benefits are negotiated, and this might account for the wage gap of $10,580 observed in our study when comparing workers with early onset disabilities to those without. It is possible that the effort of discussing accommodations might compromise time directed toward negotiating salaries or other sources of institutional support. Despite not finding statistical significance, STEM workers with disabilities acquired later in life earned on average $7,577 less per year compared to people without disabilities. Even if we accounted for full-time employment in our analysis, the fact that only 57.1% of this group reported having a full-time job suggests that acquiring a disability related to aging might result in these workers having to take on part-time positions or being unable to secure full-time jobs, ultimately leading to lower wages. Disparities in salaries between STEM workers with early onset and without disabilities were larger in the subset of those working at academic institutions than in the total sample of STEM workers. It is possible that differential allocation of grant funding or other forms of compensation could be an additional contributor to salary disparities throughout careers in academia. Even if we did not find statistically significant differences in the representation of people with disabilities across categories of reception of federal funding, previous studies have shown an imbalance in research grant success for investigators with disabilities.(3) Additionally, advancing to higher faculty ranks are key opportunities for salary raises, and the lower observed proportion of people with early onset disabilities in higher faculty ranks and leadership roles might play an important role in explaining these wage gaps. Similarly, the lower representation of workers with disabilities in the achievement of academic career milestones might be driven by unequal access to research skill building tools, such as networking and collaboration opportunities.(19)

The issue of underrepresentation of people with disabilities in the STEM workforce and in higher academic positions should be examined taking into account that 27% of the U.S. population reported having a disability in 2019.(1) This low representation could be the compounded result of a series of systematic obstacles faced by this group throughout the entire STEM educational pipeline: inadequate K-12 preparation, low expectations from teachers or faculty, insufficient mentoring, lack of familiarity of STEM teachers with specialized accommodations, and inaccessible institutions.(16, 18, 20) Consequently, students with disabilities have lower enrollment rates in STEM majors at 4-year institutions,(20) and only 9.1% of all graduates awarded doctorate degrees in 2019 reported having disabilities.(2)

Studies have highlighted that institutional environments that are inhospitable to people with disabilities lead to the attrition of talented students and workers in STEM fields,(21, 22) deepening the already existing representation gaps. Other authors have identified attitudinal and institutional barriers surrounding the issue of accommodations in higher education and STEM workplaces. On the one hand, the provision of accommodations strongly depends on the willingness of faculty and employers to fulfill these requests,(22) and evidence has shown how consistently fewer accommodations are offered in STEM fields in comparison to non-STEM fields.(23) On the other hand, bureaucratic institutional environments hamper individuals’ productivity by requiring excessive amounts of paperwork to ultimately receive accommodations, often deterring individuals from requesting services or even disclosing their disabilities status.(24) Unnecessarily cumbersome processes for assessing reasonable accommodations not only constitute an unfair burden to individuals with disabilities, but also contradict the Americans with Disabilities Act’s (ADA) purpose of embracing a social model of disability.(25) Therefore, institutional-level policies regulating disability support services must be strengthened beyond meeting the low bar of making accommodations available, the bare minimum required for ADA compliance, and should aim at the effective and timely provision of these services. Disability scholars have posited that the lack of accountability in the provision of reasonable accommodations warrants a transition to models where requests are handled by centralized institutional agencies formed by lawyers and advisors, as opposed to diffusing this responsibility among multiple departments.(21)

Even if it is well-known that students and workers with identities at the intersection of disability, race, and gender, are at a particular disadvantage for STEM opportunities,(5, 22, 26, 27) disability inclusion is not consistently integrated in dialogues pertaining diversity in STEM.(24) Evidence shows that efforts aimed at inclusion of women and underrepresented minorities in higher education have had positive results over the decades, resulting in higher representation in STEM.*(2)* Therefore, it is possible that systematically including disability issues in government-led initiatives, consolidating efforts, and setting measurable outcomes for these programs could result in reducing the representation and salary gaps for STEM workers with disabilities. Increasing representation might result in an adequate supply of role models, career champions and mentors that could help students and early career investigators with disabilities identify with STEM and later advocate for equal working conditions.(18)

As a strength, our study relies on the largest survey of U.S. doctorate recipients. Although doctorate recipients with early onset disabilities skewed younger, lower wages and underrepresentation in academic leadership roles cannot be attributed to age or longevity in the workforce. First, this group had a similar age distribution as compared to the no disability group even prior to covariate balancing. Second, in all groups a quarter of doctorate recipients were 45-54 years of age, which is likely an adequate age range to achieve full professional development. Third, differences in salaries were ascertained after balancing groups by computing propensity scores stratifying by age, and all percentages for representation in academic leadership roles were age-standardized.

Our results should be interpreted in the context of limitations. The data did not allow for analyses by disability type, and this survey uses a definition of disability developed by the Census Bureau that does not include people with psychological disabilities or learning disabilities and behavioral disorders.(28) This misclassification likely results in underestimates of the examined career and salary outcomes. Additionally, we were not able to link data on individuals across cycles of the SDR, which would have helped us shed light upon doctorate recipients’ career trajectories from a longitudinal angle. Since the SDR 2019 includes doctorate recipients that graduated between 1973 and 2017, our results are subject to period and cohort effects. Annual salaries reported by participants did not include bonuses, which are important forms of compensation for academic workers. While previous reports used working at least 30 hours per week as the threshold for defining full-time jobs, the data only allowed us to use the cut point of working at least 36 hours per week.(29). Lastly, our analysis is restricted to people who are still active in the STEM workforce, excluding 36.7% of the total SDR 2019 sample (e.g. participants who dropped out of the labor market) and introducing the potential for survivor bias.

We have documented lower wages and lower representation in STEM and academic leadership roles for doctorate recipients with disabilities. To better understand these inequities, further analyses are warranted disentangling the relationship between individual career pathways and metrics of salary and representation. Institutional compliance with ADA regulations does not translate into inclusion or equity for STEM doctorate workers with disabilities. Additional measures are needed, ranging from targeted efforts to increase participation of people with disabilities throughout STEM career pathways, to more effective provision of accommodations in higher education and workplaces for faculty and staff. Structural transformations are required to foster institutional environments where all stakeholders understand that disability services are an issue of human rights, shifting the language from accommodations to inclusion. Regarding scientific and technological development, people with disabilities are a national asset whose potential cannot be underutilized. Lower representation and salaries for people with disabilities are unjust and are not only the end result of societal structures but are also mechanisms that further place people with disabilities at a disadvantage when it comes to achieving their full professional potential, wellbeing, and participation in society.

## Data Availability

SDR 2019 public-use data are available at https://ncsesdata.nsf.gov/datadownload/.

## Acknowledgments

We would like to thank the National Science Foundation for their support with inquiries regarding SDR 2019 data.

## Authors contributions

Conceptualization: FC, ES, BKS

Methodology: FC, ES, VV, BKS

Software: FC

Formal analysis: FC, ES, JD, VV, BKS

Data Curation: FC

Writing - Original Draft: FC

Writing - Review & Editing: ES, JD, VV, BKS

Visualization: FC, ES

Supervision: BKS

---

**S1 Fig.**
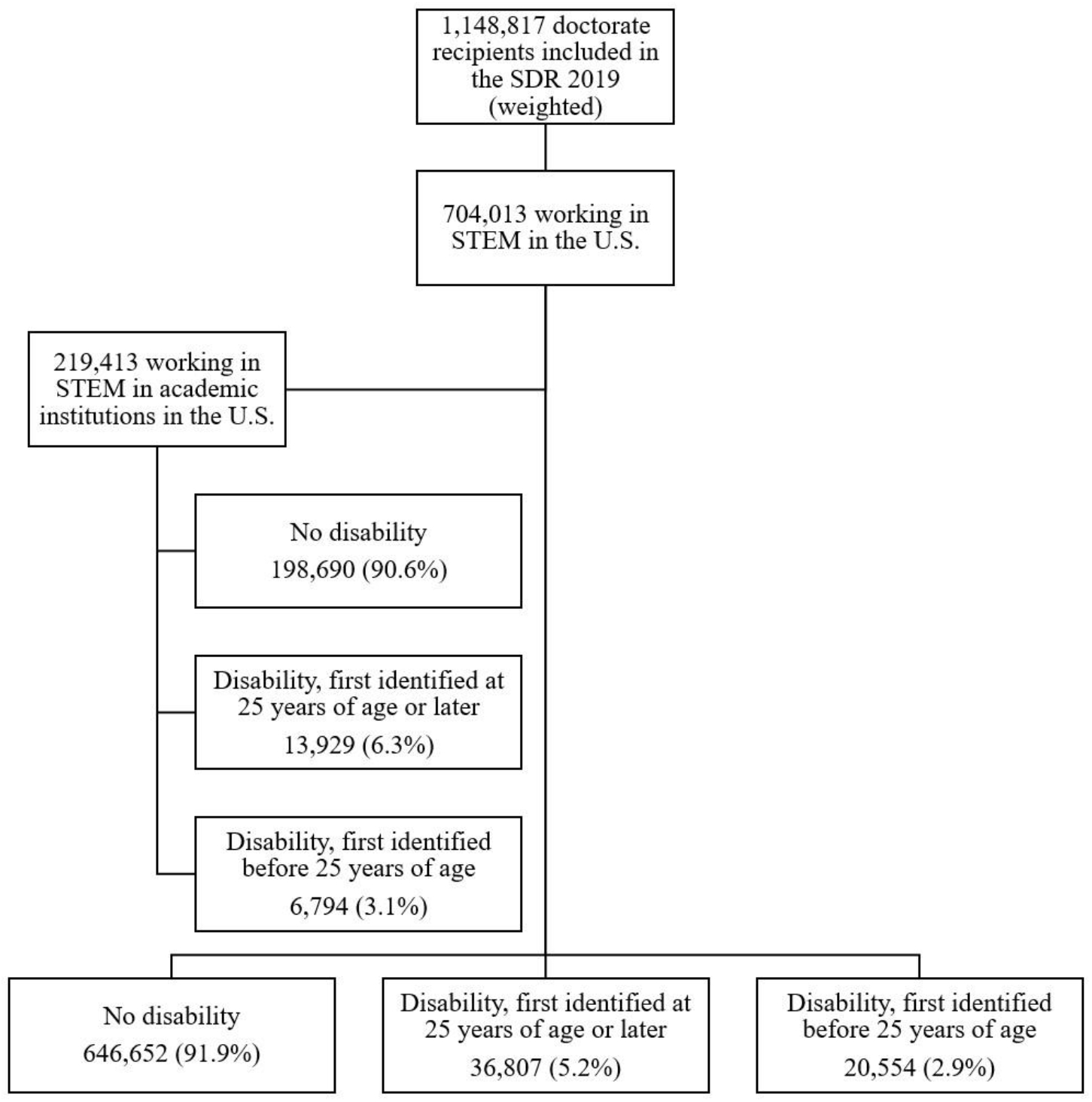
Flow diagram for study participants. Participants were currently employed in a STEM (science, technology, engineering, and mathematics) field, living in the U.S., and working for an employer based in the U.S. Academic institutions were defined as postsecondary educational institutions (4-year colleges or universities, medical schools, university-affiliated research institutes) where the following were available: faculty ranks, tenure, and academic positions (e.g. Deans or Presidents).

**S2 Fig.**
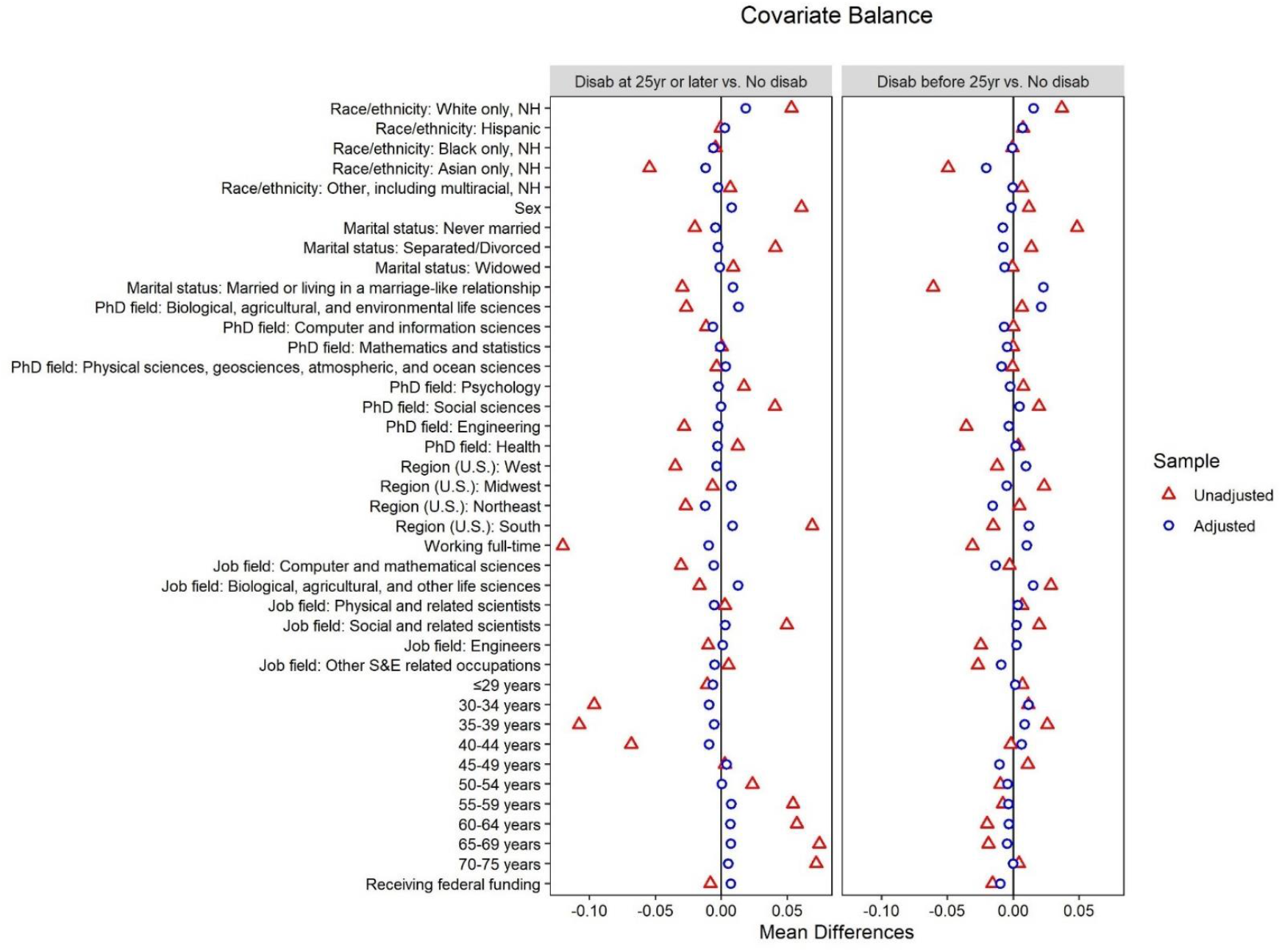
Summary of covariate balance across pairwise comparisons (late onset vs. no disability, early onset vs. no disability) for all STEM workers. Covariates included for propensity score balancing were: sex, age, race/ethnicity, marital status, degree field, job field, working full-time, receiving federal funding to support their work, region of the U.S. where the employer is located. Propensity scores were computed across strata of age (≤34 years, 35-44 years, 45-54 years, 55-64 years, ≥65 years).

**S3 Fig.**
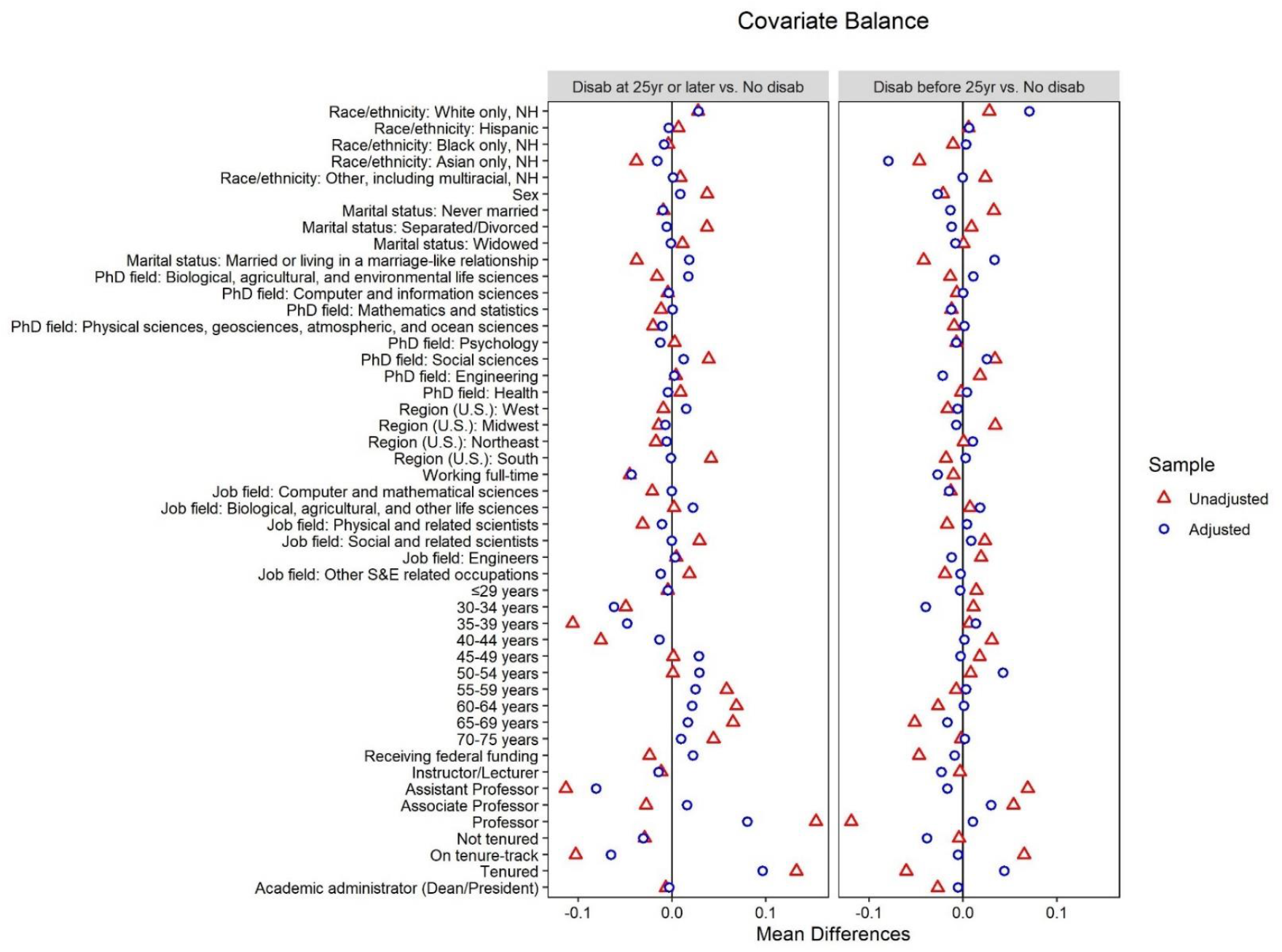
Summary of covariate balance across pairwise comparisons (late onset vs. no disability, early onset vs. no disability) in the subset of participants working in STEM at academic institutions. Covariates included for propensity score balancing were: sex, age, race/ethnicity, marital status, degree field, job field, working full-time, receiving federal funding to support their work, region of the U.S. where the employer is located, tenure status, faculty rank, being an academic administrator (Dean/President). Propensity scores were computed across strata of age (≤34 years, 35-44 years, 45-54 years, 55-64 years, ≥65 years).

**S4 Fig.**
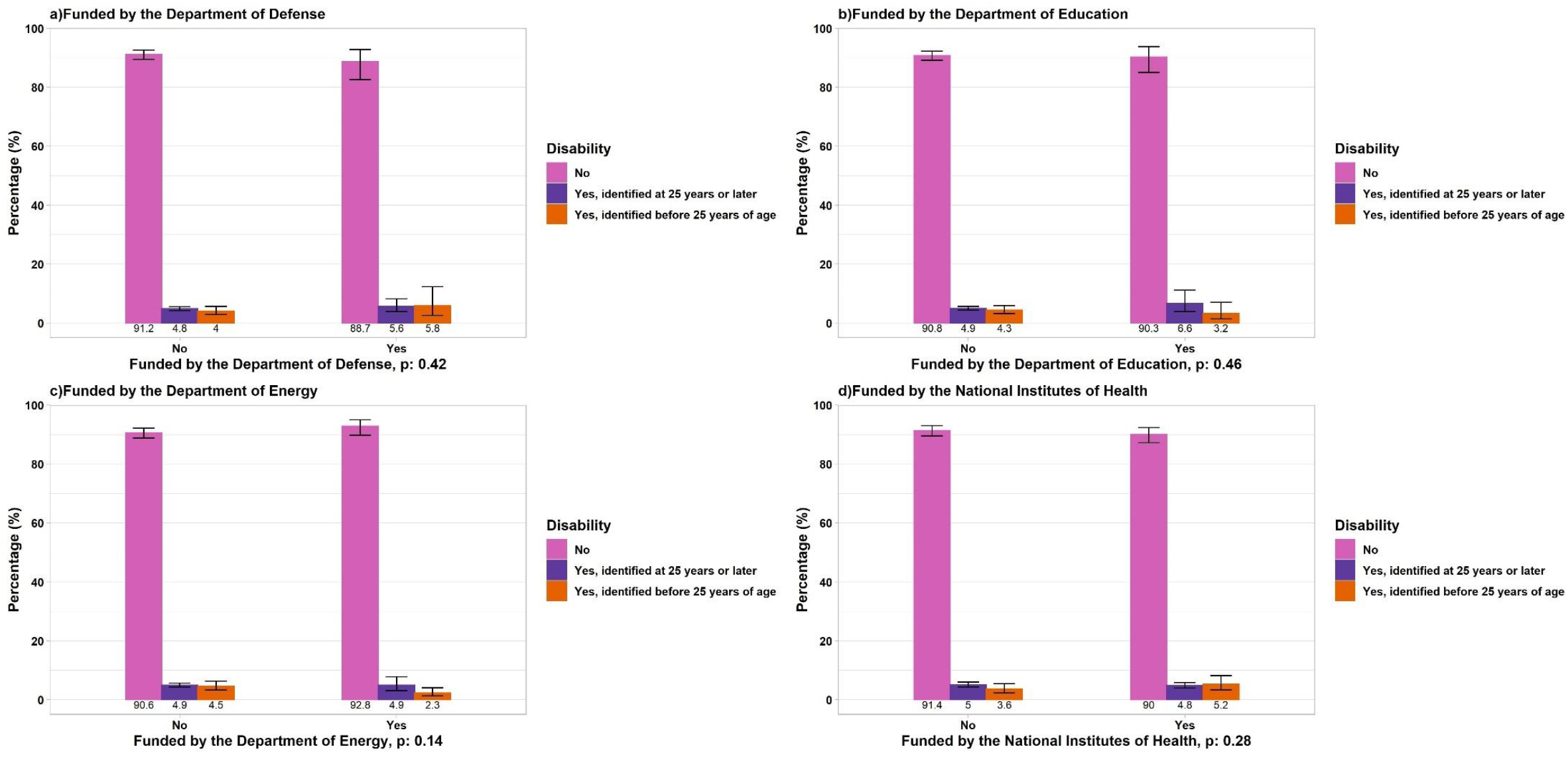
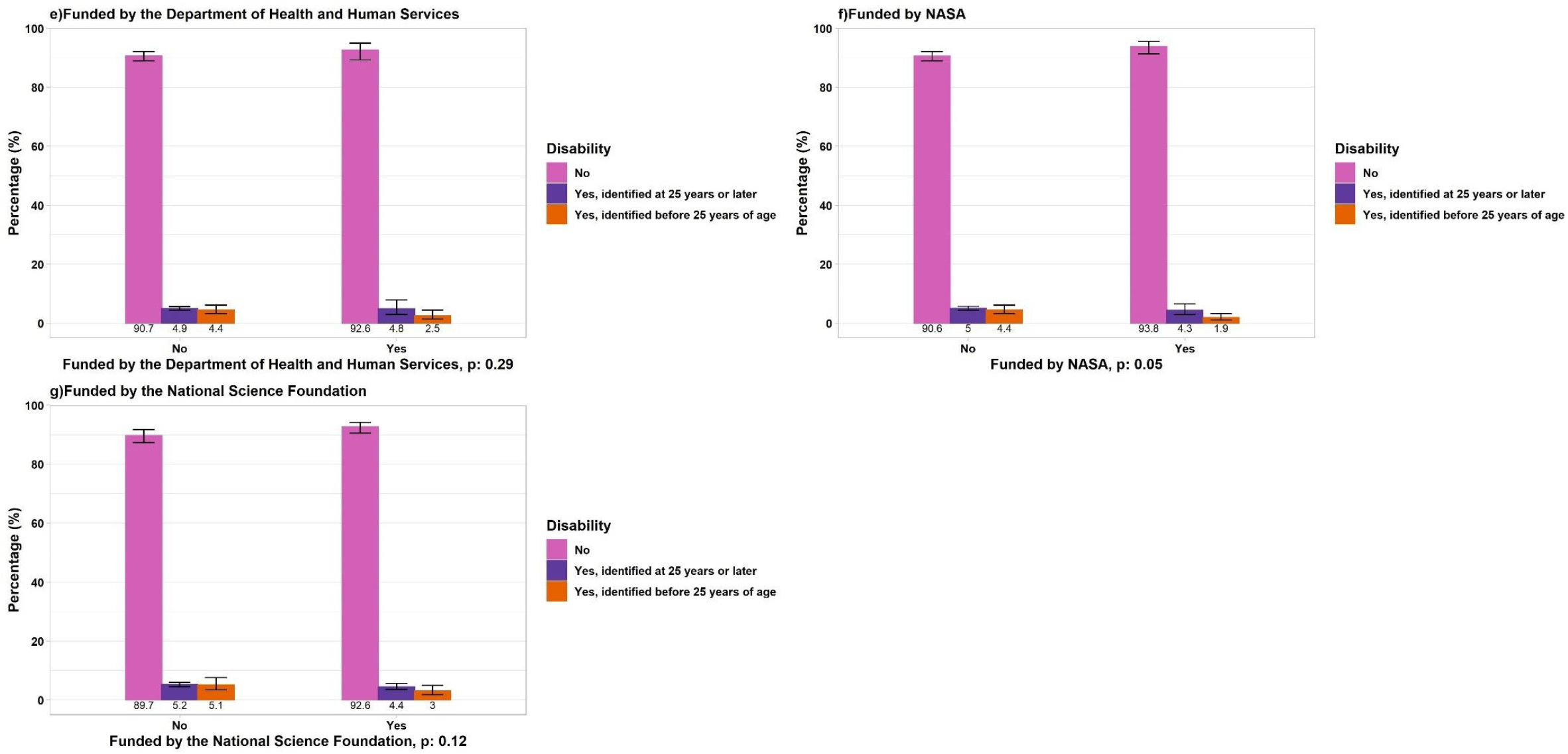
Representation (weighted, age-standardized proportions) of doctorate recipients with disabilities by type of U.S. government agency funding their work or research (grants or contracts). a) Funded by the Department of Defense, b) Funded by the Department of Education, c) Funded by the Department of Energy, d) Funded by the National Institutes of Health, e) Funded by the Department of Health and Human Services, f) Funded by NASA, g) Funded by the National Science Foundation. Other: Participants identified as “Other races including multiracial individuals, non-Hispanic” For each panel, the total denominator is the number of doctorate recipients working in STEM at academic institutions (N=219,413).

